# Development of prediction model for trauma assessment using electronic medical records

**DOI:** 10.1101/2020.08.18.20176180

**Authors:** Kentaro Ogura, Tadahiro Goto, Toru Shirakawa, Tomohiro Sonoo, Hidehiko Nakano, Kensuke Nakamura

## Abstract

Abbreviated Injury Score (AIS) and Injury Severity Score (ISS) scores are used to measure the severity of trauma patients in the emergency department, but they have several problems such as a calculation complexity. In this study, we developed a mortality prediction model of trauma patients using the data from electronic medical records and compared it with a model using AIS/ISS scores. This is a prognostic study using the data of patients who were admitted to Hitachi General Hospital Emergency and Critical Care Center from April 2018 to March 2019. The features were age, sex, vital signs, and clinical diagnoses, and the outcome was in-hospital death. Of 337 eligible patients, 11 died during the hospitalization. The predictive performance of our model was comparable to that of the AIS/ISS scores model (AUC 0.912 vs 0.961). Clinical diagnoses were important in predicting the mortality rate. Our study suggests that a trauma severity index calculated by the predicting model using information from electronic medical records might replace AIS/ISS score.

## INTRODUCTION

The initial assessment of trauma patients in the emergency department (ED) is essential for decision making and predicting their prognosis. To date, Abbreviated Injury Score (AIS), Injury Severity Score (ISS), and Revised Trauma Score (RTS) are widely used as indicators of patient severity. In addition, the Trauma and Injury Severity Score (TRISS) has also been used to calculate the life-saving potential or survival rate of trauma patients[1–2].

These scoring systems are clinically useful but have several concerns, such as calculation complexity and subjective measurements. To address these issues, studies suggest that the use of electronic medical records for predicting the prognosis of patients with trauma. For example, a prediction model using the International Classification of Diseases (ICD) had better prediction ability compared to a model using ISS[3]. However, because physicians’ diagnoses do not include pathophysiological indicators, the prognostic ability should be enhanced by the combination use of physicians’ diagnoses at the ED and vital signs as physiological indicators.

In this study, instead of using conventional indicators such as AIS and ISS scores to predict deaths, we developed a model to predict in-hospital death in patients hospitalized for trauma using patient demographics, vital signs, and the physician’s clinical diagnosis in the ED.

## METHODS

### Study design and setting

This is a prognostic study using data of patients who were admitted to Hitachi General Hospital Emergency and Critical Care Center from April 2018 to March 2019. The hospital is a tertiary emergency medical institution in Hitachi City, Ibaraki Prefecture, Japan, which covers emergency patients in the northern part of the prefecture, with 6,500 emergency transportations and a response rate of 99.9% in 2019. The Hitachi General Hospital has 18 ICUs and six cardiac care units (CCUs). The study protocol was approved by the Ethics Committee of Hitachi General Hospital, and they waived informed consent as to the nature of the retrospective design.

### Study population

We included both children and adult patients with trauma. We excluded patients with out-of-hospital cardiac arrest (OHCA).

### Candidate predictors

The candidate predictors were age, sex, vital signs (JCS: Japan Coma Scale, SBP: Systolic Blood Pressure, DBP: Diastolic Blood Pressure, PR: Pulse Rate, RR: Respiration Rate, SpO2: Saturation of percutaneous oxygen, O2: total Oxygen concentration, BT: Body Temperature) before and after arriving at the hospital, and physicians’ clinical diagnoses in the ED. These data were collected through the Next Stage ER system (TXP Medical Co. Ltd., Tokyo, Japan). The system provides prespecified forms, such as vital signs, chief complaints, past medical history, physical assessments, and clinical diagnoses, to be filled by the physicians and the nurses. As a result, the inputted clinical information is saved as reliable structured data[4].

### Outcome measure

The primary outcome was in-hospital death.

### Statistical analysis

All analyses were performed with Python (version 3.7.4; https://www.python.org/).

### Feature selection and pre-processing

Vital signs were performed for logarithmic transformation and normalization. Each diagnosis was set to 1 if the patient had the corresponding disease name and otherwise set to 0. The total values of each trauma part and the whole were also created as additional variables. To build prediction models, we preliminary built models using several combinations of these variables (**see Supplemental materials Appendix A**). For ED diagnoses, we categorized the diagnoses according to the traumatic body region. The details of this pre-processing were shown in **Supplemental materials Appendix B**.

### Training and test set

Since the small number of deaths relative to the total number of data, we used SMOTE approaches to against class imbalance (**Supplemental materials Appendix C**)[5, 6, 7]. In this study, we tried all combinations of oversampling limitations (10%, 20%, and 30% of no / survivors) and weighting the error function or not for each combination of features used in the model.

### Model building

We adopted the gradient-boosting model (GBM), which has been reported in various studies[8, 9]. GBM is a decision tree based algorithm that aims to improve accuracy by continuously learning a large number of decision trees. LightGBM is an implementation of GBM, which is known for its fast computation time and high predictive performance [10]. The Bayesian Optimization algorithm is adopted to adjust the parameters in LightGBM. This algorithm, in comparison to a total search method such as grid search, searches around the parameters that are considered to have high prediction performance [11]. This enables us to find the optimal global solution more efficiently. We examined both the use of trauma disease name information as a categorical variable and the non-use of it.

### Performance measures

The prediction models were evaluated by sensitivity, specificity, and area under the receiver operator curve (AUROC). In this study, as a comparison, we also evaluated the prediction ability of AIS and ISS scores for the trauma categories used in the proposed method for comparison: head and neck, face, chest, extremities and pelvis, abdominal and pelvic organs, and body surface, with a 1–6 value of trauma severity.

### Model selection by cross-validation

Because of the class-imbalance, the validation score with 5-fold cross-validation was used in this study to compare the prediction performance of the model (**Supplemental materials Appendix D**). The validation AUROC score of each model was calculated five times, and the model with the highest average score was determined as the best model.

### Model interpretability

In this study, the SHAP score was employed to assess the importance of features in the prediction model [12]. In each sample’s neighborhood, a linear model that approximates the mortality prediction model is created, and each feature’s coefficients are treated as SHAP scores representing importance. To create one approximate linear model for a single sample, the number of combinations of SHAP values will be created for the number of data. Besides, the larger the absolute value of SHAP value, the more significant the impact of the feature on mortality prediction. Importance has been used for feature evaluation of decision tree-based models.

## RESULTS

From April 2018 to March 2019, there were 344 patients hospitalized for trauma. Of these, we excluded seven with OHCA, and the remaining 337 patients were eligible for the analysis. The patient characteristics are shown in **Table 1**. Median age was 71 years (interquartile range 54–81) and 40.1% were men. Of 337 patients,11 patients (3.3%) died in the hospital. There were no significant differences in patient characteristics between the survival and death groups.

Table 1

**Table 1**
| Patient characteristics | The number of patients<br>with missing<br>information, n(%) | Survive<br>(n = 326) | Death<br>(n = 11) | P value |
| --- | --- | --- | --- | --- |
| age, mean (SD) | 0 | 65.4 (20.8) | 72.3 (28.7) | 0.449 |
| sex, n (%) |  |  |  |  |
| male | 0 | 130 (39.9) | 5 (45.5) | 0.760 |
| female | 0 | 196 (60.1) | 6 (54.5) |  |
| Traumatic disease count, mean<br>(STD) | 0 | 3.7 (3.8) | 4.1 (3.0) | 0.642 |
| JCS before arrival., mean (SD) | 32 | 0.8 (1.8) | 3.3 (3.3) | 0.123 |
| sBP before arrival, mean (SD) | 41 | 143.9 (33.9) | 148.4 (46.5) | 0.806 |
| dBp before arrival, mean (SD) | 50 | 84.0 (22.2) | 96.2 (8.5) | 0.058 |
| RR before arrival., mean (SD) | 94 | 21.6 (4.3) | 21.5 (3.0) | 0.971 |
| SpO2 before arrival, mean (SD) | 88 | 96.2 (3.9) | 94.0 (5.2) | 0.463 |
| BT before arrival, mean (SD) | 117 | 36.9 (2.6) | 37.6 (1.9) | 0.443 |
| sBP after arrival, mean (SD) | 56 | 147.5 (30.7) | 138.5 (52.5) | 0.694 |
| dBp after arrival, mean (SD) | 57 | 89.4 (45.8) | 89.8 (33.5) | 0.974 |
| RR after arrival, mean (SD) | 83 | 20.2 (4.4) | 23.8 (7.1) | 0.318 |
| SpO2 after arrival, mean (SD) | 73 | 97.7 (2.5) | 94.7 (3.9) | 0.122 |
| BT after arrival, mean (SD) | 80 | 36.7 (0.9) | 36.5 (1.5) | 0.856 |

Among the combinations of each predictor, the predictors used in the best model were age, gender, traumatic disease name, the total values of each trauma part and the whole, logarithmic pre-hospital vitals, and original post-hospital vitals. Of these, only sex was treated as a categorical variable. Oversampling with SMOTE was performed until the deaths were 30% of the survivors, and the error function of the model was weighted. The validation score was AUROC = 0.912 (0.815 – 0.960). The validation score of the AIS/ISS model was AUROC = 0.961(0.899 – 0.974), and the detail of the predictor was shown in **Supplemental materials Appendix E**. Of the five results calculated in selecting the best model, the each ROC curve of one representative result of which AUROC score was nearest to the average was shown in **Figure 1**.

**Figure 1.**
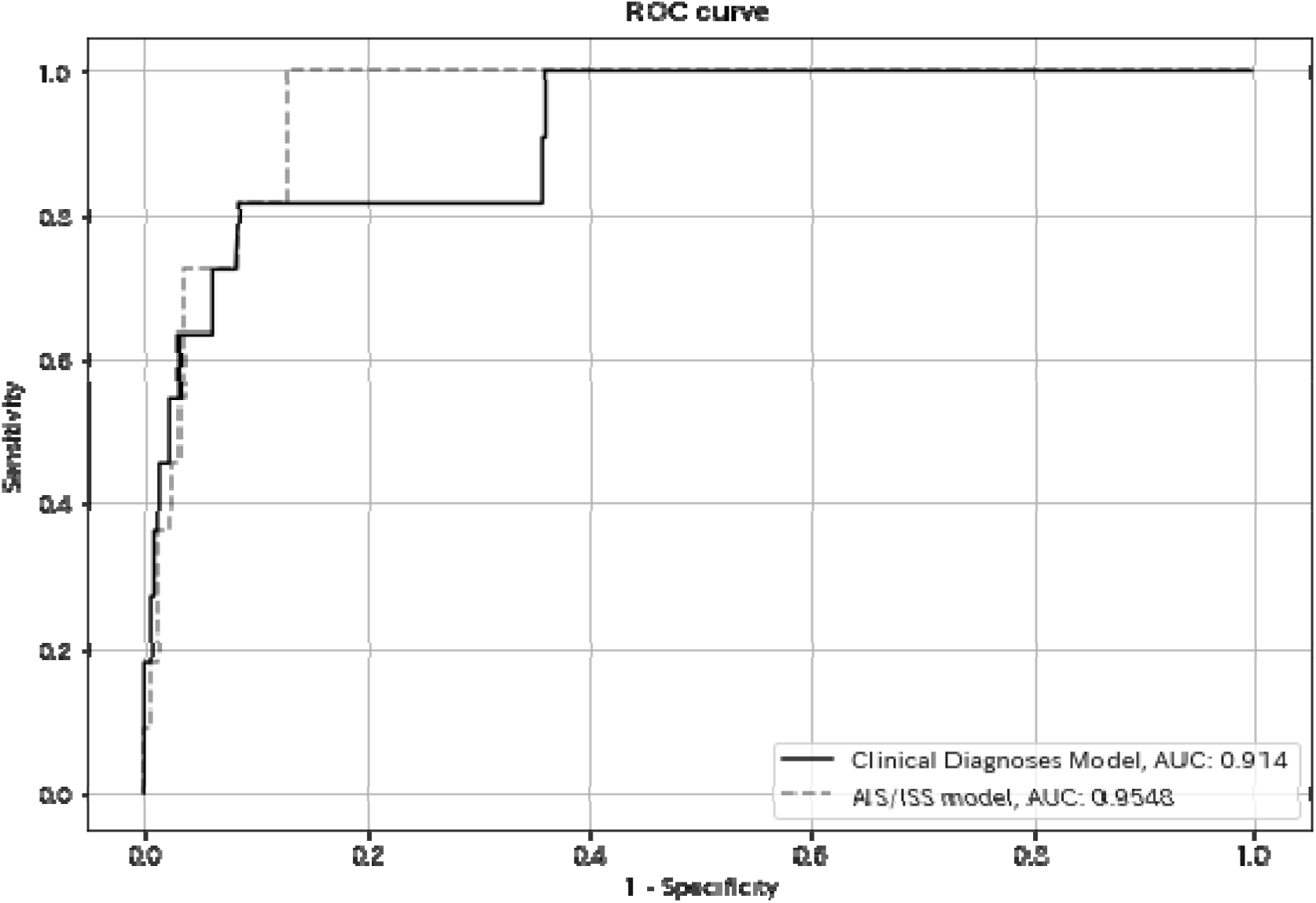
Of the five results calculated in selecting the best model, the ROC curve of one representative result of which AUROC score was nearest to the average. The ROC curve of the model using Clinical Diagnoses is drawn by the solid line, while the one of the AIS/ISS model is drawn by the dotted line.

In our model, Head and neck – hematoma, Body Temperature, Blood Pressure, Saturation of percutaneous oxygen, Head and neck – contusion, Age, and PR had a significant impact on the model output. The SHAP values of every feature for every sample were plotted in **Figure 2**, and the color represents the feature value; red high, blue low. It showed that patients who had hematoma or contusion on their head and neck, low Body Temperature, or low Blood Pressure were predicted to have a high risk of death. While the plot in **Figure 2** sorted features by the sum of SHAP value magnitudes over all samples, the bar plot in **Figure 3** clearly showed the mean absolute value of the SHAP values for each feature. **Figure 4** and **Figure 5** showed the SHAP values calculated on the AIS/ISS model. While Body Temperature had the highest impact on the death prediction, ISS score was the second most significant feature.

**Figure 2.**
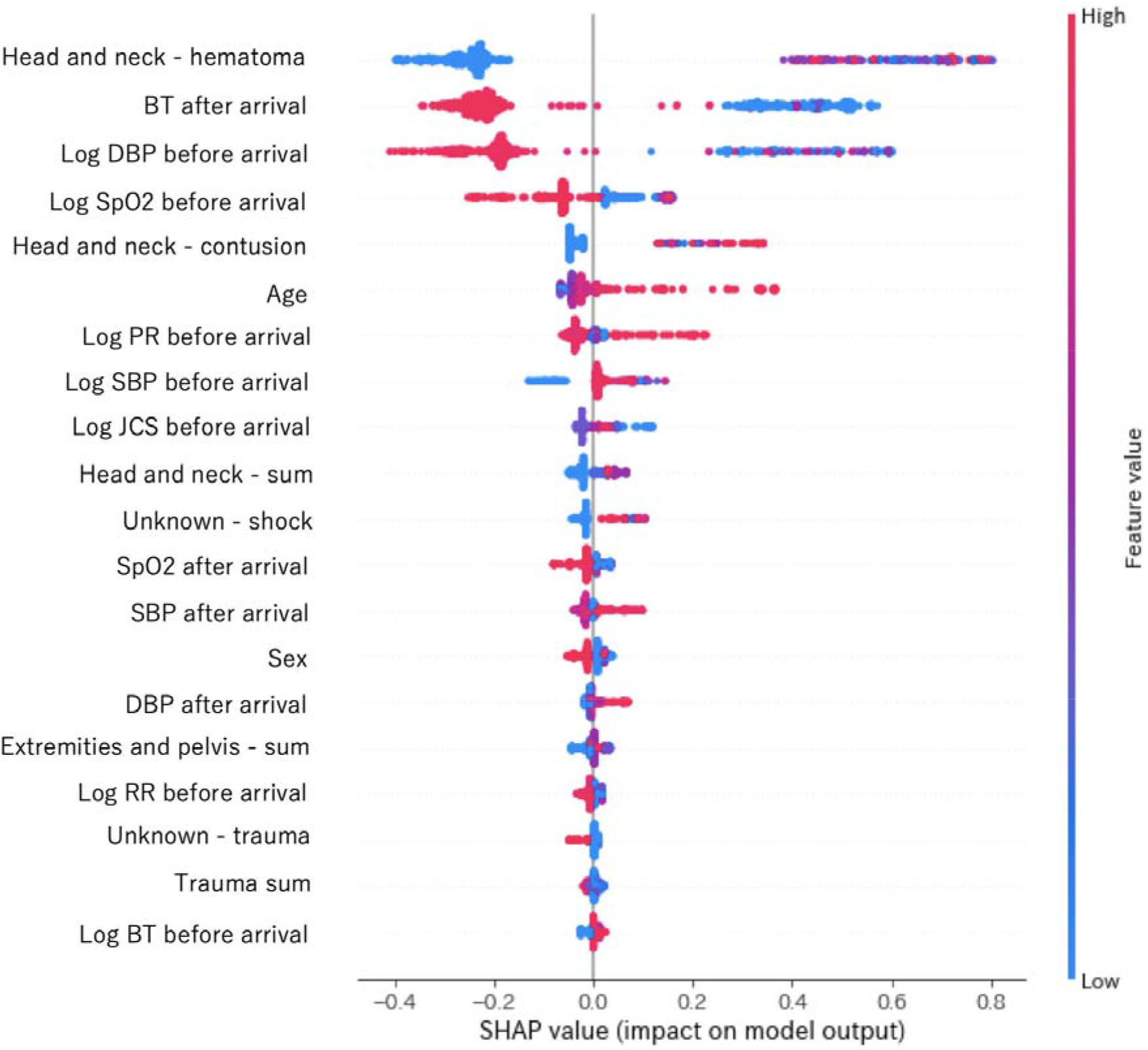
For the Clinical Diagnoses model, the SHAP values of every feature for every sample were plotted, and the color represents the feature value; red high, blue low. The plot sorted features by the sum of SHAP value magnitudes over all samples.

**Figure 3.**
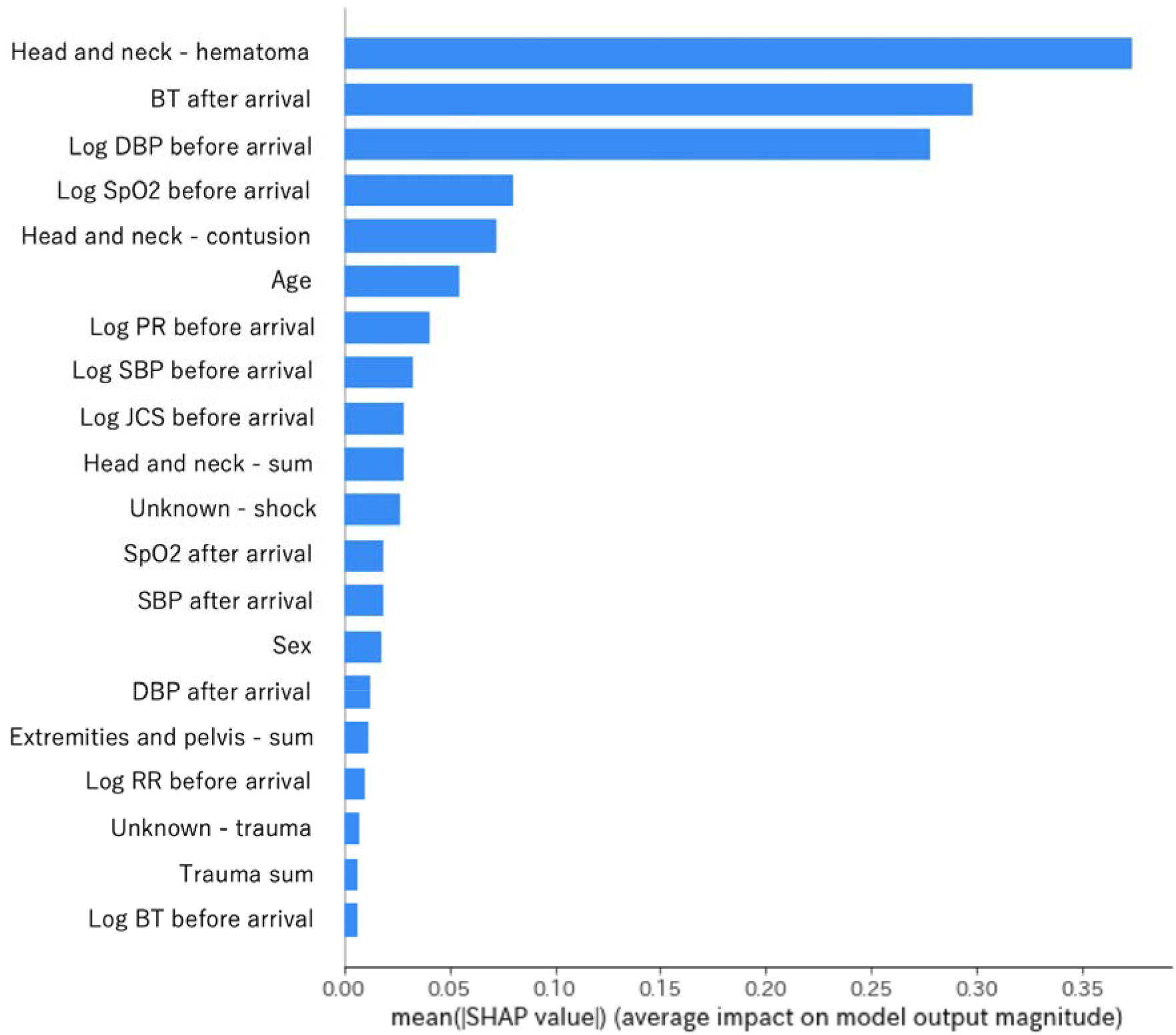
For the Clinical Diagnoses model, the bar plot showed the mean absolute value of the SHAP values for each feature, and sorted features by that value.

**Figure 4.**
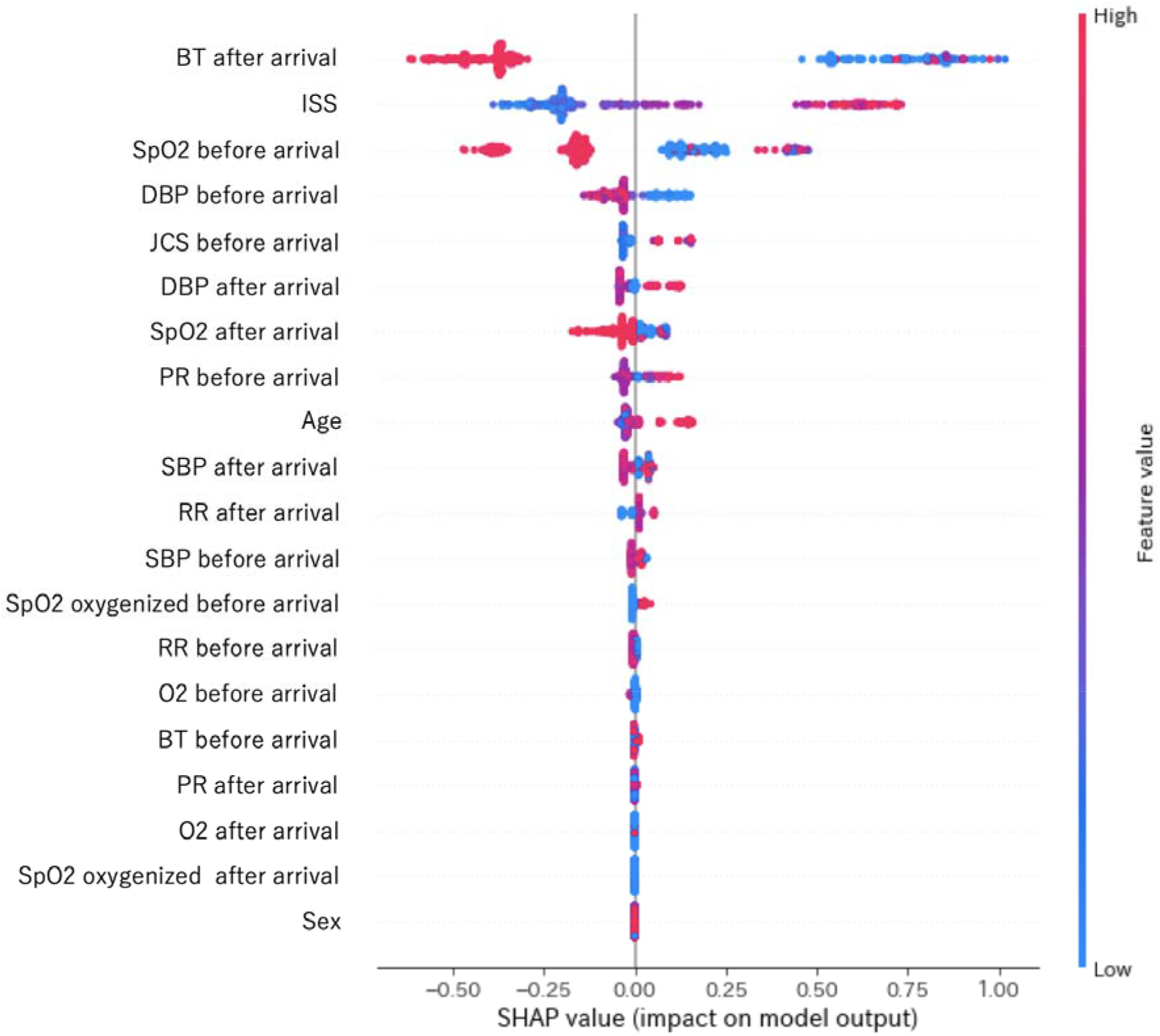
The plot values is based on the AIS/ISS model.

**Figure 5.**
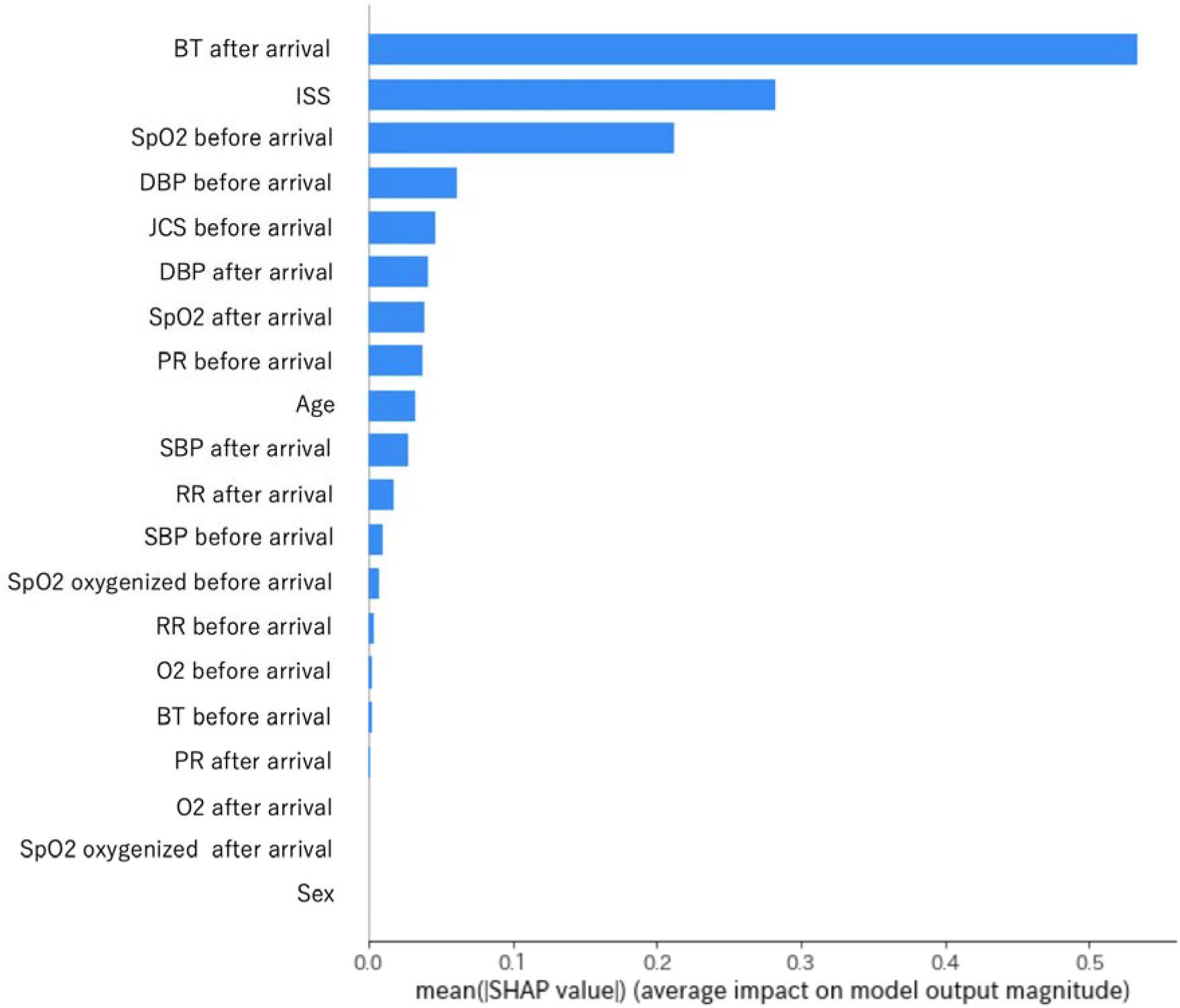
The bar values is based on the AIS/ISS model.

## DISCUSSION

Based on the data of 337 patients hospitalized for trauma, the newly developed prediction models using clinical diagnosis and vital signs showed comparable discrimination ability in predicting inhospital mortality to the prediction models using AIS and ISS. The importance of vital signs such as body temperature, blood pressure, pulse rate, conscious state, respiratory rate, and oxygen saturation has been emphasized among the features incorporated into the model. When diagnoses were added as a feature, “Head and neck – hematoma,” “Head and neck – contusion” were highly significant, followed by the total number of trauma in head and neck, “Unknown – shock”.The high importance of these head and neck traumas and shocks is consistent with the clinical perspective[13]. These results suggest that for trauma patients, a standardized list of trauma names with appropriate processing techniques, combined with vital signs, may be able to predict prognosis as well as the current ISS score.

The gold standard to date for predicting prognosis for trauma patients is a study using the National Trauma Data Bank in the United States, which used AIS scores [14]. The AUC for mortality prediction in this study was 0.83, which is the gold standard to date. However, the complicated ISS and AIS calculations are essential, and the characteristics and management of trauma patients in the U.S. and Japan are different, making it challenging to apply the U.S. predictive model to Japan as is. Similar to our study, Osler et al. in the United States reported that the prediction accuracy of a trauma mortality prediction model created by calculating the importance of the disease name itself exceeded that of a trauma mortality prediction model created using the ISS [15]. Despite the differences in patients and medical backgrounds between the U.S. and Japan, it is noteworthy and plausible that the prediction model created by calculating the importance of the trauma name itself is at least as good as the prediction model using AIS and ISS. Although the interpretation of the difference in AUC is controversial because the number of deaths in this study is significantly small, at least this study will provide an essential basis for building a predictive model of trauma patients in Japan in the future based on standard medical records of trauma rather than being as labor-intensive as in the past.

The reason for the comparable prediction performance of the AIS and ISS scores in this study is that the use of somewhat more comprehensive trauma names resulted in less information loss and allowed the model to incorporate information relevant to the prediction of death. It is also reported that the conventional method of calculating the ISS score is to multiply the coefficients by each parameter linearly, but the ISS score with separately adjusted coefficients showed an increase in the accuracy of mortality prediction compared to the current ISS score [14]. In the present study, body temperature, which is not included in the conventional gold standard mortality prediction using AIS / RTS scores, was among the second-best or best choice parameters in the models, while contributions such as respiratory rate were not necessarily high. This suggests the possible limitations of the methodology of loading by linear coefficients on parameters, both concerning the classification of trauma names and concerning the treatment of vital signs.

### Potential Limitation

There are several potential limitations in this study. First, the relatively small sample size limits the model building processing. However, we used the SMOTE approach to add data on deaths like the new sample data rather than merely increasing the number of deaths. Second, because this is a singlecenter study, the generalizability of the findings may be limited by regional and institutional characteristics (e.g., differences in treatment methods for trauma, characteristics of patients receiving treatment in an aging region). Lastly, the impact of the exclusionary patient criteria may reduce the AUC score of the predicting model. This is because patients with cardiopulmonary arrest at the time of presentation were excluded in this study, and they tend to show a high mortality rate.

## CONCLUSIONS

In this study, for patients with trauma, we found that the model for predicting in-hospital mortality using electronic medical records has comparable prediction ability compared to those using time-consuming AIS and ISS scores.

## Data Availability

The datasets generated and/or analyzed during the current study are not publicly available due to no IRB approval of data sharing.

